# Countering Neural Activity Drift: Sustained Long-term Seizure Prediction Using an Evolutionary Machine-Learning Framework on Continuous Intracranial EEG

**DOI:** 10.64898/2026.09.14.26359203

**Authors:** Justo Montoya-Gálvez, Karla Ivankovic, Marzieh Nazari, Alessandro Principe, Rodrigo Rocamora

## Abstract

**Objective:** Seizure prediction in drug-resistant epilepsy remains a major biomedical challenge. Traditional machine learning approaches rely heavily on segmented offline testing, which suffers from artificial class rebalancing, hides neural activity drift over time, and severely inflates performance estimates. This study introduces an online evolutionary framework designed for realistic brain-computer interface validation and addresses performance degradation caused by neural drift.

**Methods:** We present the first publicly available, continuous long-term stereoelectroencephalography dataset for seizure prediction, tracking 16 patients across 664.9 hours of data and 121 seizures. An offline combinatorial analysis evaluated pipeline decisions (referencing, frequency bands, functional connectivity metrics, and classifiers). The highest-performing offline elements, namely monopolar referencing, cross-correlation-based connectivity matrices, and Random Forests classifiers, were evaluated under realistic class imbalances in a pseudo-prospective online framework. Candidate models (N = 1,000 per patient) were evolved across continuous streams, while a data-driven approach targeted patient-specific epileptogenic networks for input channel reduction.

**Results:** Transitioning from offline to online evaluation demonstrated a prominent performance drop, with mean AUROC degrading from 90% to 57.9%, confirming offline metrics conceal temporal neural drift. However, our online evolutionary framework identified models achieving complete event-level seizure prediction in 75% of patients (and all but one seizure in 87.5%). Restricting inputs to the estimated epileptogenic network identified using our previously proposed approach reduced implanted contact requirements by 79.12% while providing superior predictive performance over clinically resected areas.

**Significance:** This work demonstrates that actionable, deterministic seizure prediction is achievable when paired with modern computational capability. Because 1,000 candidate models represent a conservative proof-of-concept search budget, scaling search spaces in production environments can further expand optimal interictal sampling and prediction yields. By providing both a continuous benchmark dataset and a standardized online evaluation protocol, this framework offers a foundation for self-adapting closed-loop devices capable of post-event retraining and long-term deployment in clinical neuromodulatory

**Key Points:**

- First publicly available continuous long-term iEEG dataset for seizure prediction, with the largest patient and channel count to date.
- The proposed framework predicted all seizures in 75% of patients, and all but one seizure in 87.5% of patients.
- Online evaluation reveals substantial performance overestimation in offline testing, highlighting the need for realistic BCI validation.
- Epileptogenic network-based channel selection achieves comparable predictive performance with around 79% fewer implanted contacts.
- Classifier choice and functional connectivity metric are the dominant drivers of seizure prediction performance across the pipeline.

## 1. Introduction

Epilepsy affects approximately 50 million people worldwide ^1^, with around one third of patients developing drug-resistant epilepsy (DRE) ^2–3^, posing a significant clinical challenge. While resective surgery targeting the epileptogenic zone is the most established treatment option for DRE ^4^, it has long-term failure rates of 40–50% ^5^, and surgical success depends critically on accurate identification of the epileptogenic network (EN), for which no gold-standard methodology exists.

As an alternative to surgery, neuromodulation devices emerged. The most prominent example is the Responsive Neuro Stimulation ® system (RNS; NeuroPace Inc.), presented to detect seizures and enact an electrical stimulation using the implanted electrodes ^6^. The RNS improved on the Vagal Nerve Stimulation (VNS), but even the best reports show that its efficacy falls behind the surgical treatment ^7^. Predictive closed-loop systems (CLSs), conceptualised as brain-computer interfaces (BCIs), aim to continuously monitor EEG or intracranial EEG (iEEG), predict seizure onset, and deliver neuromodulatory interventions only when needed ^8^, unlike earlier systems that stimulate independently of the underlying brain activity. Within this context, seizure prediction can be formulated as a brain-state classification task, in which the objective is to distinguish the preictal state from other epileptic brain states ^9^.

The first prospective, long-term, human clinical trial employing invasive EEG for seizure prediction, the NeuroVista Seizure Advisory System ^10^, demonstrated that seizure prediction is feasible. Notably, the device relied exclusively on subdural electrode strips, which are more susceptible to artefacts and do not provide access to deep brain structures. Some subjects had perfect predictions, however the overall results moved the NeuroVista team to promote a Kaggle event to improve the prediction algorithms ^11^. The results from the competition, coupled with the perception that reliable neuromodulatory prediction may not yet be feasible ^12^, progressively shifted the academic research interest toward seizure forecasting rather than seizure prediction. However, most patients and their caregivers prefer seizure prediction horizons (SPHs) shorter than 24 hours, which usually are not contemplated by forecast systems ^13^. Today, modern data-center infrastructure, high-performance computing (HPC), and cloud-based artificial intelligence (AI) processing, could allow the field to re-engage directly with short-horizon, deterministic seizure prediction

We argue that achieving reliable prediction requires moving beyond model architecture optimization to address dataset curation and pipeline standardization. Our recent review highlighted the scarcity of publicly available epilepsy datasets as a major bottleneck in seizure prediction research. Moreover, public datasets are overwhelmingly composed of brief, pre-segmented snippets ^11^ with artificially rebalanced interictal/preictal class ratios ^7^. This forces most studies to rely on limited offline segmented analyses and risk model overfitting and performance inflation ^14^, as they are aimed to be deployed under long-term, highly imbalanced conditions. Few studies have systematically examined this offline-to-online transition ^11^, despite evidence that online assessment of concept drift can improve seizure prediction performance ^15^.

Additionally, the effects of EEG-specific preprocessing decisions on seizure prediction performance remain underexplored. Although preprocessing procedures such as signal cleaning, normalisation, outlier handling, and oversampling improve performance ^16^, evidence is lacking on referencing schemes, band filtering, functional connectivity (FC) metrics, dimensionality reduction, and electrode target selection. Existing evidence from motor, cognitive, and clinical neuroscience suggests that these choices substantially influence downstream connectivity and classification results ^17–20^. Similarly, dimensionality reduction approaches based on leading eigenvectors have shown promise for EEG state classification tasks ^21^.

Together, these findings highlight the need for a standardised yet flexible framework capable of handling inter-patient variability while balancing predictive performance and computational efficiency for clinically viable seizure prediction devices. In this context, channel reduction strategies may be particularly relevant, as reducing EEG channels has been associated with improved sensitivity ^22^.

To address these challenges, we introduce the first publicly available, continuous long-term stereo-EEG (SEEG) dataset specifically curated for continuous seizure prediction benchmark testing. Then, we perform an exhaustive combinatorial analysis of signal preprocessing choices, connectivity metrics, and classifiers under offline conditions. Lastly, we introduce a computationally scalable, online evolutionary framework designed for prospective BCI deployment.

## 2. Materials & Methods

The study was divided into two stages to compare offline and online evaluation settings (Fig. 1). In the first stage, classifiers were trained and tested using limited, non-continuous data segments to identify candidate models for seizure prediction. In the second stage, candidate models were evaluated on longer continuous recordings to better approximate real-world conditions.

**Figure 1.**
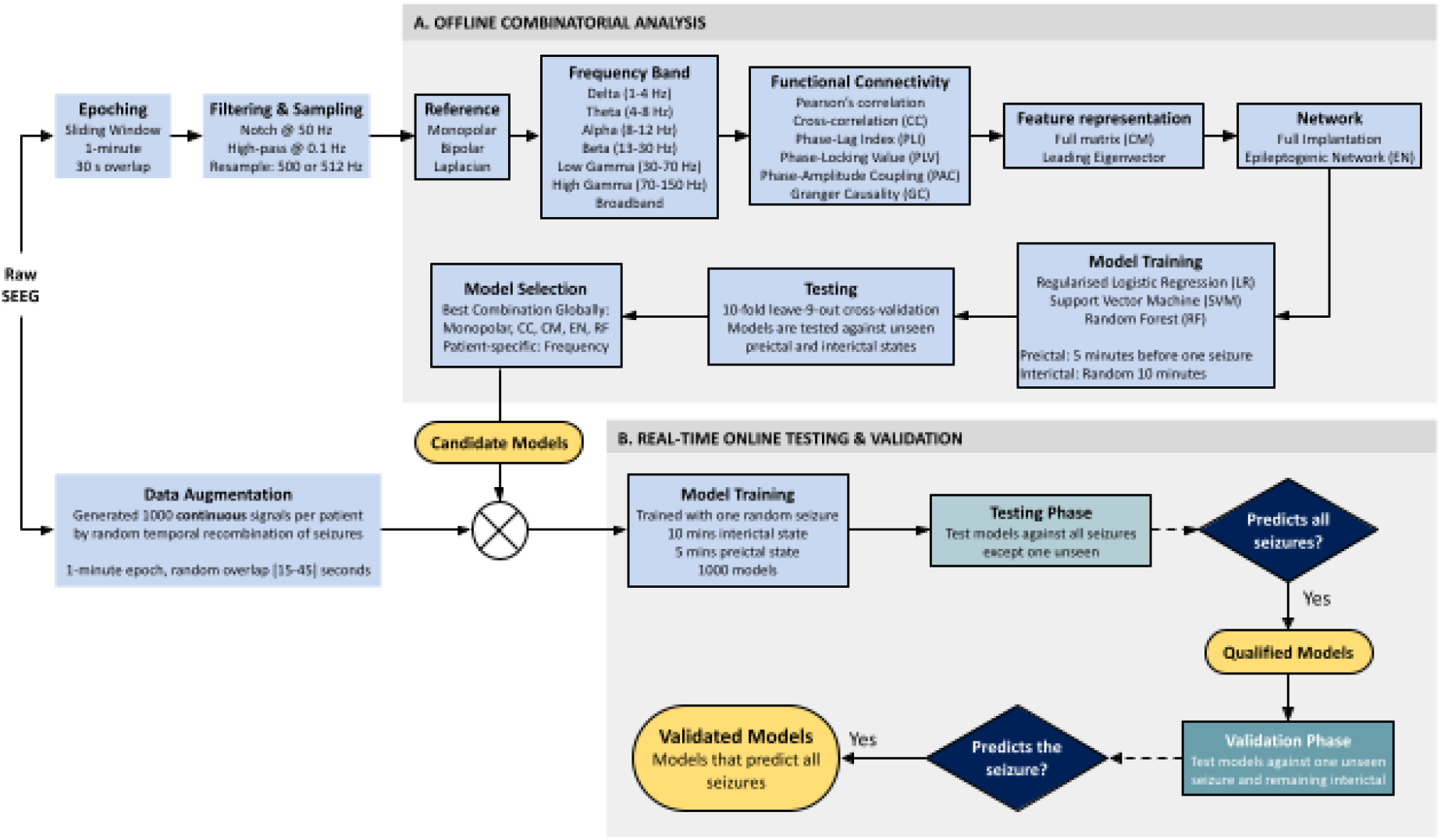
Overview of the seizure prediction framework. The diagram summarises the complete workflow followed in this study. Rectangles represent processing stages, diamonds represent decision points, the circle with a cross denotes a summing junction, and the stadium shape indicates workflow termination. Block A illustrates the offline analysis pipeline, including signal preprocessing, feature extraction, model selection, and model training. Block B illustrates the real-time online testing and validation framework used to identify candidate models capable of predicting all seizures. Diagram elements without an outline correspond to steps performed once, whereas outlined elements indicate stages repeated across multiple configurations in parallel. For example, SEEG signals were processed independently using monopolar, bipolar, and Laplacian references, and models were trained and evaluated across multiple combinations of frequency bands, connectivity measures, feature representations, and classifier types. SEEG, stereoelectroencephalography; EN, epileptogenic network; PLV, phase-locking value; PLI, phase-lag index; PAC, phase-amplitude coupling; LR, Logistic Regression; SVM, Support Vector Machine; RF, Random Forest.

### 2.1 Patient Cohort

We retrospectively analysed SEEG recordings from patients with DRE who underwent presurgical evaluation at the Epilepsy Monitoring Unit of Hospital del Mar (Barcelona, Spain) between 2013 and 2019.

Inclusion criteria required non-palliative resective surgery, focal epilepsy, at least three years of postoperative follow-up, and a minimum of two spontaneous seizures recorded during presurgical SEEG monitoring. The follow-up criterion was chosen to ensure reliable classification of surgical outcome, as seizure relapse risk decreases substantially after this period ^23^. A detailed breakdown of inclusion and exclusion criteria is provided in Table S1.

### 2.2 Electrode Implantation & Data Acquisition

All patients underwent stereotactic implantation of depth electrodes for presurgical evaluation at Hospital del Mar, with clinically determined implantation schemes independent of this study. Intracranial electrodes (Dixi Médical, Besançon, France; details in Appendix S1) were implanted using the ROSA robotic system.

SEEG recordings were acquired using a clinical monitoring system (XLTEK, Natus Medical) at sampling rates between 250 and 2,048 Hz. Continuous recordings were used when available, and missing segments were filled to preserve temporal continuity. Recordings affected by stimulation were excluded unless acquired at least 24 hours after the last stimulation. Seizure onsets and offsets were visually annotated and classified as clinical or subclinical. Data were exported in EDF+ format using a monopolar reference montage ^24^.

### 2.3 Epoching Strategy

The preictal state was defined as the 5 minutes preceding clinical seizure onset, with 60-second epochs with 30-seconds overlap sampled within this window (Appendix S2). The interictal class included all data excluding ictal periods, preictal intervals, and the 60 minutes preceding seizures. The interictal class size was set to twice the number of preictal epochs to mitigate class imbalance.

### 2.4 Referencing

Three referencing schemes were evaluated: monopolar, bipolar, and Laplacian. Monopolar used the original extracerebral reference, bipolar computed adjacent contact differences, and Laplacian enhanced local activity by attenuating common components (Appendix S3).

### 2.5 Artifacts Removal & Sampling Rate Normalization

Power-line interference was removed using notch filters at 50 Hz and its harmonics with a 5 Hz stopband (Appendix S4). A 0.1 Hz Butterworth high-pass filter at 0.1 Hz was then applied to reduce slow drifts.

Recordings were harmonised in sampling rate. Signals acquired at 250 Hz were upsampled to 500 Hz, while those at 1024 Hz and 2048 Hz were downsampled to 512 Hz.

### 2.6 Filtering

Signals were bandpass filtered into canonical frequency bands delta (1–4 Hz), theta (4–8 Hz), alpha (8–12 Hz), beta (13–30 Hz), low gamma (30–70 Hz), and high gamma (70–150 Hz) using a 2nd-order Butterworth filter. Each band, including broadband activity, was analysed independently.

### 2.7 Functional Connectivity

FC was estimated using Pearson correlation, cross-correlation (CC), Phase-Lock Value (PLV), Phase-lag Index (PLI), Phase-Amplitude Coupling (PAC), and Granger causality (GC). For each epoch, a normalized N x N connectivity matrix (CM) was computed across nodes and analysed separately for each referencing scheme and frequency band. PAC was restricted to broadband signals, while PLV was not computed on broadband data.

### 2.8 Feature representation of the connectivity matrix

To assess latent structure in CMs, spectral decomposition was performed and only the leading eigenvector was retained. Both full and reduced representations were then evaluated.

### 2.9 Network Size: Full Implantation vs. Selected Nodes

Two node sets were analysed as targets: the full implantation and the EN. Since seizure prediction systems are typically intended to avoid resective interventions, no ground-truth EN is available in practical applications. To address this limitation, the EN was estimated using our previously proposed machine learning–based approach for identifying putative epileptogenic networks from connectivity data ^25^.

### 2.10 Classifiers

Connectivity matrices were vectorised and used as input to three common ^26^ classifiers of increasing complexity: logistic regression (LR), support vector machine (SVM), and random forest (RF). Three baseline models were also included (majority, minority, and random stratified classifiers). The SVM produced calibrated probabilities, and class imbalance in the RF was handled using inverse-frequency weighting (Appendix S5).

### 2.11 Performance Metrics

A stratified 10-fold leave-(k−1)-out cross-validation scheme was used, preserving interictal-to-preictal ratios. Each training iteration used one fold for training and nine for testing. Only preictal data from one seizure was included in the training set per iteration. Performance was evaluated using AUROC, balanced accuracy, sensitivity, specificity, precision, and training time.

### 2.12 Model Comparison

Main-effects analysis was performed to quantify the contribution of the pipeline factors to AUROC. Due to violations of parametric assumptions, Welch’s ANOVA with Games–Howell post hoc tests was used. Dimensionality reduction effects were assessed using Kruskal–Wallis tests with Dunn’s correction. Efficiency was evaluated using two composite metrics (AUROC per training time and AUROC per node), also analysed with Kruskal–Wallis and Dunn’s tests.

### 2.13 Selection of the Best-Performing Models for Online Testing

Offline analyses were used to set the pipeline configuration for online evaluation. Monopolar referencing, CC-based connectivity, full CM representation, and EN-based node selection were retained as default settings. RF was selected as the classifier. Frequency band selection was performed individually per patient based on offline AUROC maximisation.

### 2.14 Data Augmentation for Online Testing

To better approximate long-term deployment conditions, continuous recordings (15 hours to 3 days per patient) were expanded through repeated resampling. Signals were segmented into 60-second epochs with random overlap (15–45 s), repeated 1000 times to increase data volume (Appendix S6). Epochs were labelled as interictal, preictal, ictal, subclinical, or transition.

### 2.15 Training

For each patient, 1000 classifiers were trained. Each classifier used one randomly selected seizure, with preictal CMs extracted from the 5 minutes preceding that seizure and interictal data sampled from 10 random minutes of interictal activity. Random epoch resampling ensured variability in the resulting CMs across classifiers, even when using the same seizure.

### 2.16 Online Testing

Each classifier was evaluated in two pseudo-prospective stages: an initial leave-one-seizure-out testing phase, and a validation phase under realistic class imbalance. Synthetic continuous recordings were constructed by concatenating seizure-related blocks and randomising seizure order while preserving local temporal structure (Appendix S7). Models were then applied in an online manner using a 60 s sliding window, generating preictal predictions based on RF probability thresholds and a 5-minute persistence rule.

Performance was assessed at both sample and event levels using recall and AUROC. To better capture clinically relevant dynamics, event-based metrics were prioritised.

Only models achieving perfect event-level recall during the testing phase (i.e., predicting all seizures), termed qualified models, were retained for validation. The validation stage incorporated an expanded interictal block to better reflect the class imbalance of continuous recordings. Models that also achieved perfect event-level recall during validation were considered validated seizure predictors, with their relative performance subsequently characterised using secondary metrics.

## 3. Results

### 3.1 Dataset

Of 50 screened patients, 16 met inclusion criteria (10 women, 6 men; age [mean ± SD] 46.76 ± 13.72 years; range 21–69) and were included. Patients were stratified by surgical outcome at last follow-up into a good outcome group (Engel I, n = 12; 6 women, 6 men; age 52.33 ± 15.48 years; range 31–69) and a bad outcome group (Engel II–IV, n = 4; 4 women; age 33.4 ± 9.1 years; range 21–45).

A total of 1,779 SEEG channels were analysed (104.65 ± 36.05 per patient; range 27–150), corresponding to 664.9 h of recordings (36.94 ± 12.36 per patient; range 15.07–71.95). Across all subjects, 121 seizures were identified (7.56 per patient; median 5, SD 6.54; range 2–28), with 91 in the good outcome group and 30 in the bad outcome group.

To our knowledge, this constitutes the first publicly available intracranial EEG dataset based on continuous recordings and one of the largest in terms of patients and channels, surpassed in duration only by the Kaggle American Epilepsy Society Seizure Prediction Challenge dataset (Table S2, Appendix S8).

Given that EN inference is constrained by SEEG sampling, analyses involving EN comparison were restricted to patients with good surgical outcomes, in whom EN identification is considered reliable. In this subgroup, ENs estimated using our previously proposed approach (see Methods: 2.9. Network Size) were identified in all patients, showing correspondence with resected regions, and selection of EN nodes (291 channels; 16.17 ± 9.14 per patient; range 8–36) resulted in a 79.12% reduction in implanted contacts relative to full implantation.

### 3.2 The Classifier and Connectivity Method are Primary Determinants of Prediction Performance

Welch’s ANOVA revealed significant effects across all processing stages (all p < 0.001), with classifier type and connectivity method as the main determinants of AUROC (Table S3). Classifier choice had the largest effect (η◻² ≈ 0.27), followed by connectivity method (η◻² ≈ 0.045), while other factors contributed marginal effects. Overall, performance was primarily driven by model and connectivity selection.

### 3.3 Performance Ranking of Classifiers and Connectivity Metrics via Post Hoc Analysis

Games–Howell post hoc analysis (Table S4) showed a consistent ranking of methods (all p < 0.001). RF achieved the highest AUROC (0.944), followed by LR (0.855) and SVM (0.597) (Fig. 2A–C). Among connectivity measures, CC performed best (0.887), followed by Pearson correlation (0.799) and PLV (0.791). Overall, classifier and connectivity choice dominated performance differences.

**Figure 2.**
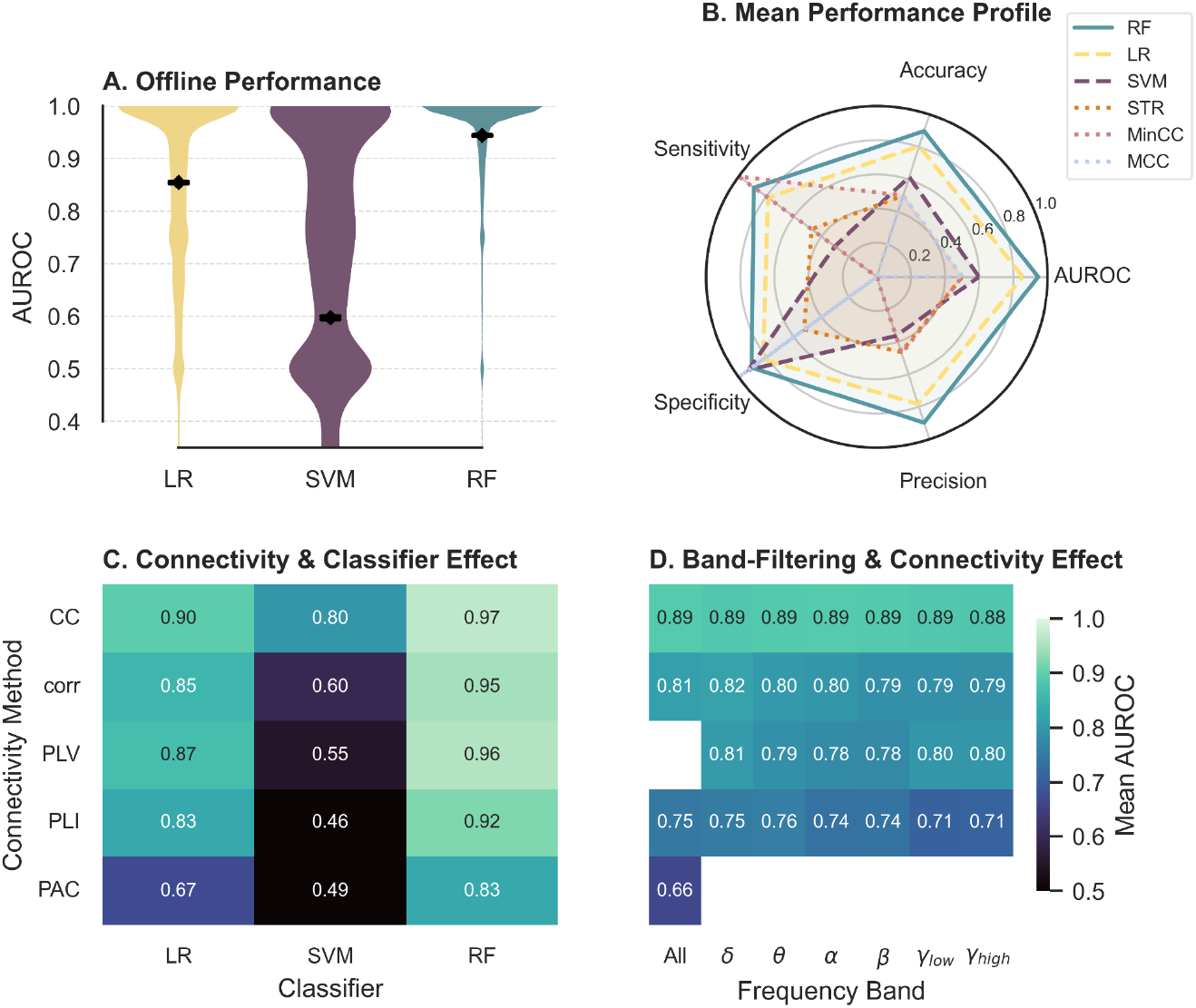
Average offline performance of the evaluated classifiers and connectivity configurations. **(A)** Violin plots showing the distribution of classifier performance, measured by area under the receiver operating characteristic curve (AUROC), for Logistic Regression (LR), Support Vector Machine (SVM), and Random Forest (RF) classifiers. Black markers indicate the mean AUROC for each classifier. **(B)** Radar plot depicting the mean performance profile of the evaluated classifiers, including AUROC, accuracy, sensitivity, specificity, and precision. Results are shown for LR, SVM, RF, random stratified (STR), minority class (MinCC), and majority class (MCC) classifiers. **(C)** Heatmap showing the mean AUROC stratified according to classifier type and connectivity method. Lighter colours indicate higher AUROC values. **(D)** Heatmap showing the mean AUROC stratified according to frequency band and connectivity method. CC, cross-correlation; corr, Pearson’s correlation; PLV, phase-locking value; PLI, phase-lag index; PAC, phase-amplitude coupling; All, broadband; δ, delta band (1-4 Hz); θ, theta band (4-8 Hz); α, alpha band (8-12 Hz); β, beta band (13-30 Hz); γ_low, low gamma band (30-70 Hz); γ_high, high gamma band (70-150 Hz).

### 3.4 Secondary Pipeline Factors: Effects of Referencing, Connectivity Representation, and Band Filtering

Reference scheme, CM representation, and band filtering showed small effects (η_p_^2^ ∼ 0.01). Monopolar referencing achieved slightly higher performance (AUROC = 0.816) than bipolar (0.790) and Laplacian (0.791), with no difference between bipolar and Laplacian (p = 0.977). Full CM representation outperformed eigenvector-based reduction (0.807 vs 0.791; p < 0.001). Band-specific performance (Fig. 2D) showed modest differences: delta yielded the highest AUROC (0.817), followed by high gamma (0.796) and low gamma (0.795), with mixed significance across comparisons (Appendix S6). The alpha band (0.800) did not differ consistently from most bands, except in selected pairwise contrasts. Overall, these findings indicate that referencing, connectivity compression, and spectral band choice have limited but measurable effects refining performance.

### 3.5 Marginal Performance Gain from Full Network Inclusion over EN

The full implantation achieved significantly higher performance (AUROC = 0.818) than the EN subset (AUROC = 0.779; p < 0.001) (Fig. 3A), although the effect size was small (Hedges’ g = 0.14).

**Figure 3.**
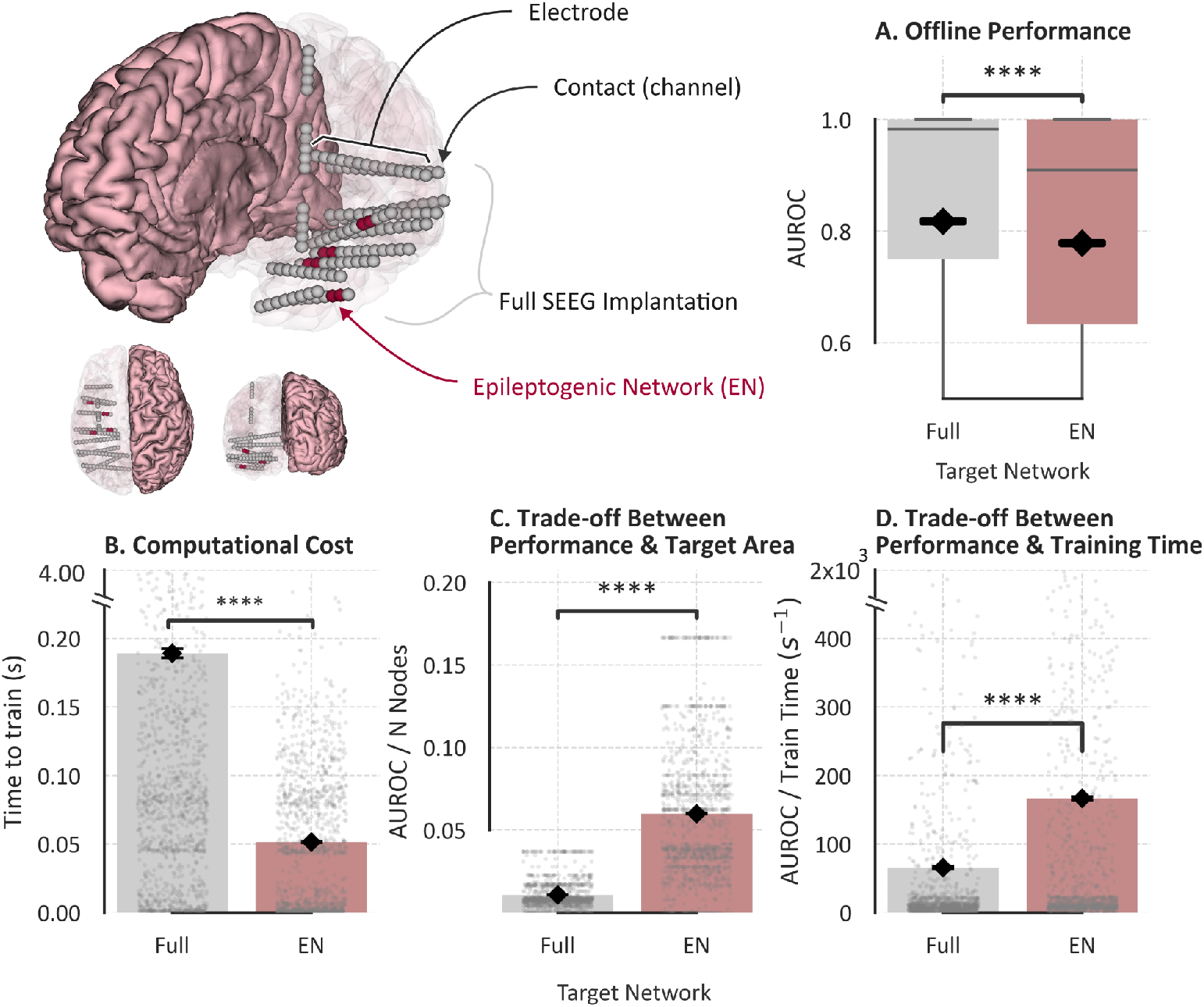
Comparison between using the full stereoelectroencephalography (SEEG) implantation and restricting the analysis to the epileptogenic network (EN). The upper-left panel shows a three-dimensional reconstruction of one patient’s brain (pink) and SEEG implantation. The opacity of the left hemisphere was reduced to visualize the implanted electrodes within the brain. Each sphere represents an electrode contact (channel). Contact size was enlarged to 5 mm for visualization purposes, although actual contacts measured 2 mm in length and 0.8 mm in diameter. Contacts belonging to the EN are highlighted in dark red, whereas the remaining contacts are shown in grey. Inferior reconstructions display superior (left) and posterior (right) views of the same implantation. **(A)** Boxplots showing the distribution of classifier performance, measured by area under the receiver operating characteristic curve (AUROC), for models using the full SEEG implantation (grey) and models restricted to the EN (red). Black markers indicate the mean AUROC for each group. There is only a modest performance gain from using the full electrode set, suggesting that restricting recordings to the EN may be sufficient to maintain strong predictive performance. **(B)** Comparison of computational cost, measured as classifier training time (seconds), between models using the full implantation and those restricted to the EN. Each dot represents an individual classifier. The y-axis was truncated to 0-0.25 s for visualization purposes, although some training times reached 4.0 s. **(C)** Trade-off between classifier performance and the number of nodes included in the target network (full implantation versus EN), expressed as AUROC per node. **(D)** Trade-off between classifier performance and computational cost, expressed as AUROC per unit of training time (s^{-1}). The y-axis was truncated to 0-500 s^{-1} for visualization purposes, although some values reached 2000 s^{-1}. Asterisks indicate statistically significant differences between groups (****, p < 0.0001, Dunn’s test with Bonferroni correction).

### 3.6 Efficiency Gains from Reduced Representations and EN-Based Networks with Favorable Performance Trade-offs

CM reduction via eigenvalue decomposition produced significant differences in computation time (Dunn’s test, p < 0.0001). The full CM required 0.181 s on average (SD: 0.455), whereas using only the leading eigenvector reduced this to 0.065 s (SD: 0.058).

Network size also significantly affected computation time (p < 0.0001; Fig. 3B). The estimated EN yielded the lowest computation times (0.051 s ± 0.053), while full implantation was highest (0.189 s ± 0.445).

Configurations based on the estimated EN also showed significantly better efficiency trade-offs between performance, number of nodes, and computation time (all p < 0.0001; Fig. 3C–D).

### 3.7 Selected Optimal Pipeline Exhibits Marked Performance Degradation in Online Evaluation

The optimal configuration across patients consisted of monopolar referencing, CC connectivity, full CM representation, and restriction to EN nodes. In the offline evaluation, this configuration achieved a mean AUROC of 0.900 (SD: 0.159; range: 0.389–1.000). However,under continuous sample-based online evaluation exposed to natural class imbalances and neural activity drift, performance decreased to 0.579 (SD: 0.149; range: 0.132–1.000).

Furthermore, sample-based evaluation overestimated performance compared to event-based metrics (Fig. S1). Accordingly, to assess whether online evolutionary model filtering could successfully counter this drift, all subsequent analyses were based exclusively on event-level metrics.

### 3.8 Online Evolutionary Model Selection, Validation Performance, and Search Space Yield

Within the initial budget of N=1000 candidate models per patient, the testing phase identified qualified models satisfying the full-prediction criterion (predicting 100% of seizures) in 14 of 16 patients (87.5%). Qualified models achieved a mean AUROC of 0.731 (SD: 0.139; range: 0.157–1.000; Fig. 4A-B), significantly outperforming disqualified models (0.449 ± 0.232; range: 0.01–0.965; Mann–Whitney U, p < 0.0001).

**Figure 4.**
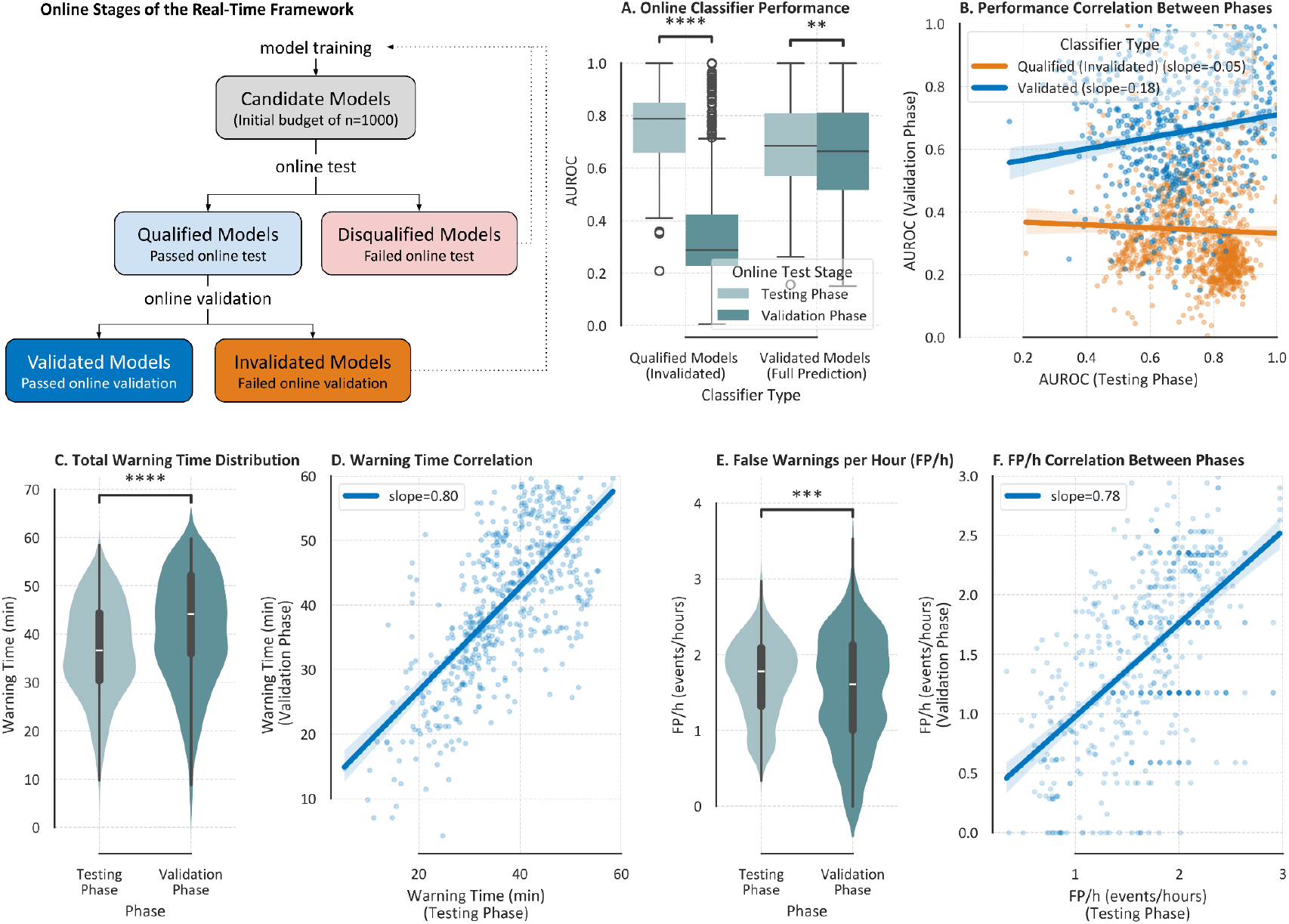
Online classifier performance and warning characteristics during the real-time framework evaluation. The upper-left panel illustrates the workflow of the online framework. Rectangles represent processing stages, diamonds represent decision points, and the stadium shape indicates workflow termination. Solid arrows indicate stages implemented in the present study, whereas dotted arrows represent recalibration steps proposed for future real-world brain-computer interface (BCI) deployment. In the current framework, classifiers failing to achieve full-prediction are discarded. (A) Boxplots showing the distribution of online performance, measured by area under the receiver operating characteristic curve (AUROC), for validated classifiers achieving full-prediction (i.e., correctly predicting all seizures) and qualified models (i.e., failing to predict at least one seizure) during the testing (light blue box) and validation (dark blue box) phases. Asterisks indicate statistically significant differences between groups (*, Mann-Whitney p < 0.0001). (B) Linear regression analysis comparing classifier AUROC between the testing and validation phases. Each point represents an individual classifier. Blue corresponds to classifiers achieving full-prediction, whereas orange corresponds to classifiers achieving partial-prediction. Regression lines and their slopes are shown for each group. (C) Violin plots showing the distribution of total warning time, defined as the cumulative duration (minutes) during which the online signal was classified as preictal, during the testing (light) and validation (dark) phases for classifiers achieving full-prediction. The violin shape represents the kernel density estimate (KDE), the white line indicates the median, the thick grey bar the interquartile range (IQR), and the thin grey lines the whiskers of the embedded boxplot. (D) Linear regression analysis comparing warning time between the testing and validation phases for classifiers achieving full-prediction. Each point represents an individual classifier. (E) Violin plots showing the distribution of false positives per hour (FP/h) during the testing (light) and validation (dark) phases for classifiers achieving full-prediction. (F) Linear regression analysis comparing FP/h between the testing and validation phases for the same classifiers. Asterisks indicate statistically significant differences between groups (, p < 0.001; ****, p < 0.0001, Mann-Whitney test).

The validated models, which successfully passed the validation phase on previously unseen continuous recordings, maintained complete event-level seizure prediction in 12 out of 16 patients (75.0%), with mean AUROC of 0.688 (SD: 0.162; range: 0.157–1.000). Crucially, validated models surviving both testing and validation represented a small fraction of the search space (mean 4.92%; median 2.65%; SD 5.37%; range 0.1–19.0%).

As all validated models achieved identical recall value, performance differentiation relied on secondary metrics. Mean warning time was 36.81 min in testing and 40.25 min in validation (Fig. 4C), with moderate agreement between phases (Pearson r = 0.8; Fig. 4D). FP/h remained similarly stable (1.69 vs 1.51 events/hour, r = 0.78; Fig. 4E-F). In contrast, SPH showed no significant correlation between testing (mean: 32.74 min; median: 20.00; SD: 40.53) and validation (mean: 37.08 min; median: 14.00; SD: 67.37).

### 3.9 Impact of Accurate EN Identification on Online Prediction Performance

To assess the importance of EN identification, online prediction was tested in patients with poor surgical outcomes using channels from the clinically resected region instead of the estimated EN channels (see Methods: 2.9. Network Size). EN-based inputs achieved significantly higher mean AUROC than resection-based inputs (0.673 vs 0.619; SD: 0.194 vs 0.162; Mann–Whitney U, p < 0.001).

Moreover, EN-based configurations consistently produced validated models capable of predicting all seizures across patients (up to 102 models in some cases), whereas resection-based inputs achieved validated models in only one patient.

### 3.10 Impact of Interictal Sampling Timing on Online Performance Variability

Because all classifiers shared identical processing pipelines, variance in online performance may be partly driven by differences in training data composition. Since preictal segments were fixed to a single seizure, performance differences could be primarily attributable to the interictal epochs used during training.

To quantify this effect, we analysed the temporal origin of interictal epochs using their mean time-to-seizure (TTS). Classifiers achieving full-prediction were mainly trained on interictal data preceding the training seizure (TTS > 0), whereas partial-prediction models were predominantly trained on post-seizure (TTS < 0) interictal data (Fig. 5). Across all classifiers, those trained on pre-seizure interictal epochs achieved higher AUROC than those trained on post-seizure epochs (0.637 vs 0.559; Mann–Whitney U, p < 0.0001).

**Figure 5.**
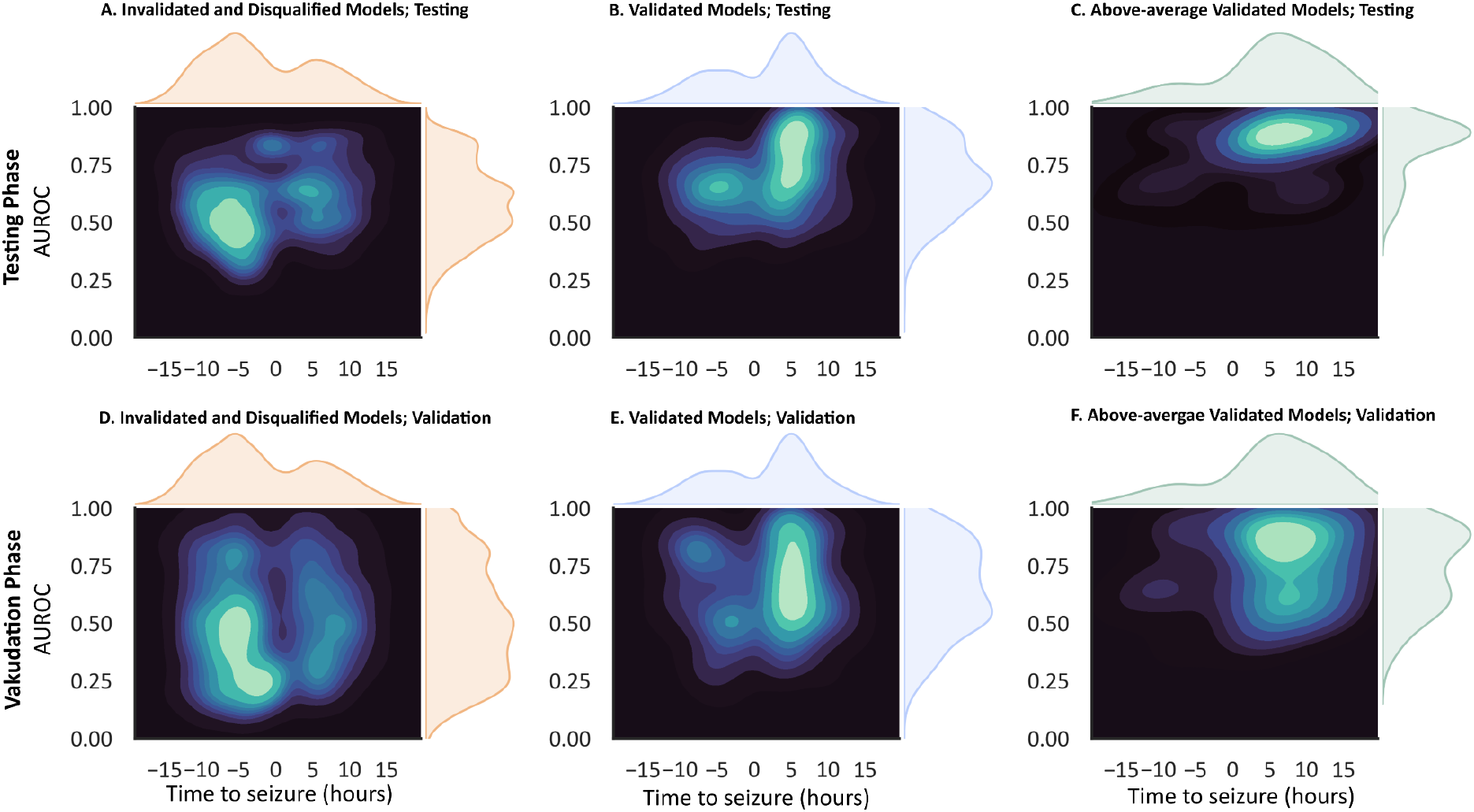
Temporal characteristics of the interictal states used for training the best-performing classifiers. **(A)** Bivariate kernel density estimate (KDE) showing the relationship between the average time to seizure (TTS, hours) of the interictal epochs used for training and the AUROC achieved by partial-prediction classifiers during the testing phase of the online framework. Positive TTS values indicate that interictal epochs were sampled, on average, before the preictal period used for training, whereas negative TTS values indicate that interictal epochs were sampled after the preictal period. Darker regions represent lower-density areas of the distribution, whereas lighter regions represent higher-density areas. Most partial-prediction classifiers were trained using interictal epochs sampled after the associated preictal state. **(B)** KDE showing the relationship between average TTS and AUROC for full-prediction classifiers during the testing phase. Most classifiers achieving full-prediction were trained using interictal epochs sampled before the associated preictal state. **(C)** KDE showing the relationship between average TTS and AUROC for above-average full-prediction classifiers, defined as the top-performing half among classifiers achieving full-prediction, during the testing phase. **(D)** KDE showing the relationship between average TTS and AUROC for partial-prediction classifiers during the validation phase of the online framework. **(E)** KDE showing the relationship between average TTS and AUROC for full-prediction classifiers during the validation phase. **(F)** KDE showing the relationship between average TTS and AUROC for above-average full-prediction classifiers during the validation phase. AUROC, area under the receiver operating characteristic curve; KDE, kernel density estimate; TTS, time to seizure.

However, under the fixed N=1000 search budget, pre-seizure interictal sampling alone was insufficient for 6 of 16 patients. In those patients, no validated models were found (i.e. achieving full prediction) when restricting interictal sampling exclusively to epochs preceding the training seizure.

## 4. Discussion

### 4.1 The Reality Gap in Continuous Seizure Prediction

Our findings reveal a substantial “reality gap” between standard offline benchmarking and continuous online evaluation. Although offline testing achieved a mean AUROC of 0.900, performance dropped to a sample-based AUROC of 0.579 during prospective evaluation on continuous recordings, consistent with previous concerns ^28^ about non-continuous evaluation ^29–30^. This degradation likely reflects the extreme class imbalance and temporal drift inherent to real-world continuous iEEG.

To address this limitation, we release, to our knowledge, the first publicly available continuous long-term SEEG dataset for seizure prediction, comprising 664.9 hours of recordings from 16 patients. The dataset provides a realistic benchmark for clinically relevant evaluation, surpasses existing public intracranial datasets in cohort size and implanted channels per patient, and offers recording durations comparable to the Kaggle American Epilepsy Society Seizure Prediction Challenge dataset ^27^. A detailed comparison with existing datasets is provided in Table S2.

### 4.2 Overcoming Neural Drift via Online Evolutionary Filtering and Production Scalability

Rather than treating neural drift as a barrier to long-term deployment, our evolutionary algorithm-inspired framework retained only qualified models that predicted all seizures during testing. Unlike previous approaches ^31–32^, the framework explicitly enforces complete event-level prediction, achieving 100% seizure prediction in 87.5% of patients during testing and maintaining this performance in 75.0% of patients on unseen validation data (Fig. 6).

**Figure 6.**
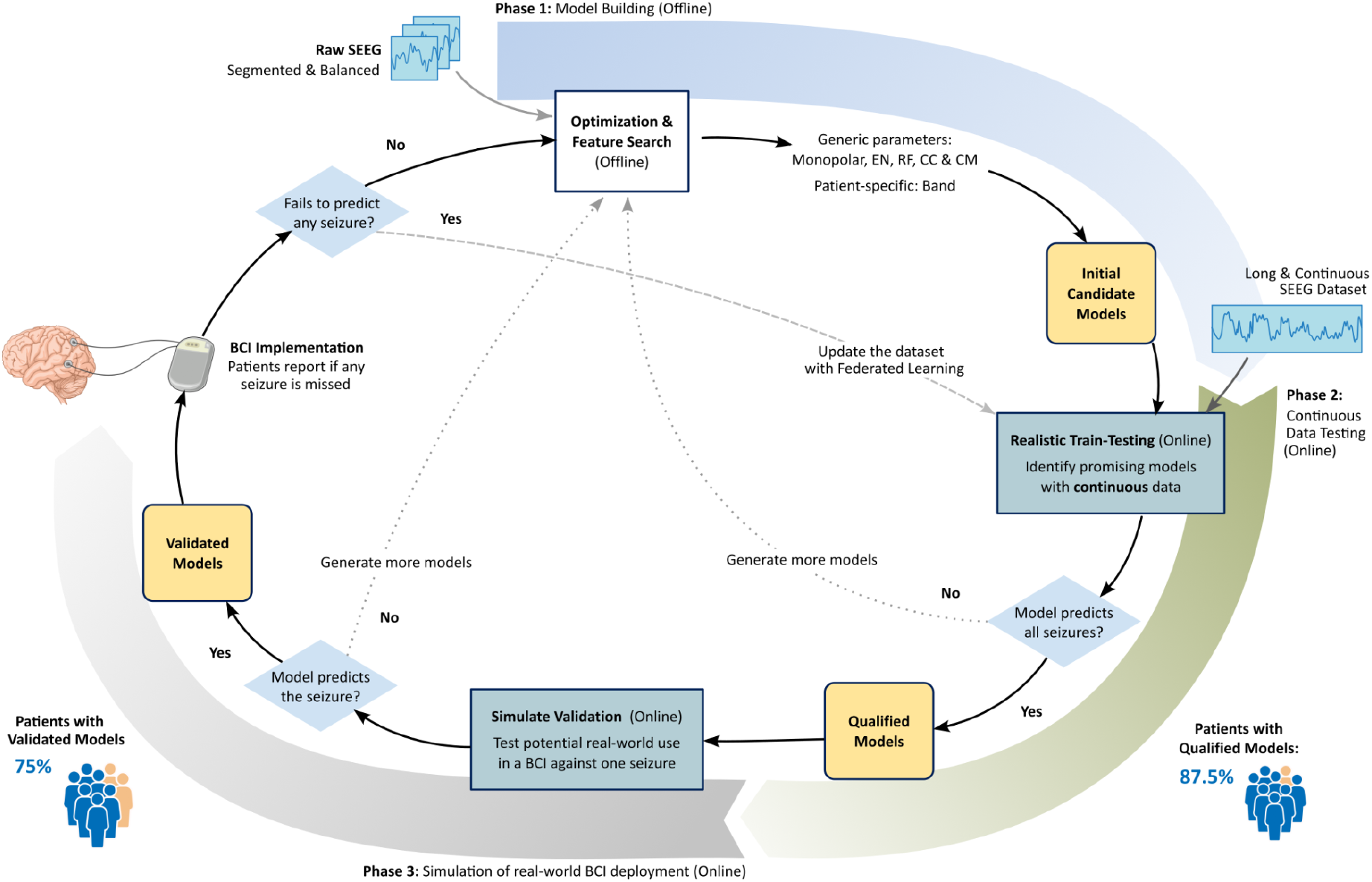
Summary of the proposed framework and its intended clinical translation. The workflow is structured into three in silico phases designed to progressively evaluate and improve seizure prediction models before potential deployment in a real-world brain-computer interface (BCI). In the first phase (Model Building; in blue), the workflow begins with offline optimisation using segmented and balanced data. Combinations of preprocessing strategies, connectivity measures, frequency bands, feature representations, and classifiers are evaluated to identify models with the highest predictive performance, referred to as Candidate Models. In the second phase (in green), Candidate Models are assessed using continuous SEEG recordings in a real-time in silico setting. To progress to the next stage, models must achieve full-prediction (i.e., correctly predict all seizures) during long-term continuous recordings, sorted by false positive rates (FP/h) in ascending order. Models meeting these criteria are designated Qualified Models and are subsequently evaluated in the third phase (in grey) using continuous recordings containing previously unseen seizures, designed to emulate prospective deployment in a real-world BCI. Models that correctly predict the unseen seizures are considered Validated Models. Validated Models may then be translated to in vivo BCI applications, where patient feedback on correctly predicted and missed seizures can be incorporated into an online learning strategy. In this scenario, the BCI would continuously acquire new data, which could be integrated through remote and federated learning to iteratively update the models, with the aim of improving generalisation to future seizures and adapting to long-term changes in brain dynamics (brain drift).

This 75.0% success rate should be considered a conservative lower bound, as it was obtained using only 1,000 candidate models per patient, of which approximately 5% satisfied both testing and validation criteria. Scaling the search to tens of thousands of candidate models with modern HPC and cloud AI infrastructures, together with continuous model updating to adapt to neural drift ^15^, could further improve performance. Our findings also show that artificially selecting interictal periods, including in curated datasets, limits the identification of models capable of predicting all seizure events, highlighting the importance of continuous recordings.

Finally, false warning time and FP/h remained highly consistent between testing and validation, suggesting stable model behaviour under realistic conditions. Although the median SPH exceeded the 5-minute target (20 min in testing and 14 min in validation), it may still be clinically relevant ^13^.

### 4.3. Mechanism of Model Resilience: Interictal Timing and Continuous Retraining

A key finding of this study is the importance of the temporal origin of interictal training data. Classifiers trained on interictal segments preceding a seizure (TTS > 0) achieved significantly better validation performance than those trained on post-seizure interictal data. This likely reflects the heterogeneity of interictal activity, where pre-seizure segments are closer to the evolving seizure trajectory, whereas post-seizure segments are influenced by postictal recovery and broader non-stationarities ^15^, consistent with cyclic seizure generation models ^47–48^. These results suggest that prioritising pre-seizure interictal sampling may improve model robustness by providing more informative negative samples.

This finding also supports an iterative post-event retraining strategy (Fig. 6), in which each new seizure recording is incorporated into a cloud-based optimisation process that continuously updates the candidate model pool, allowing the BCI to adapt to neural drift and maintain performance during long-term deployment..

### 4.4. Importance of Network Selection for Hardware-Constrained Implantables

For seizure prediction to be translated into practical implantable BCIs, input channel requirements must be compatible with hardware power and computational constraints of miniature, battery-powered implants. Our EN node estimation strategy (see Methods: 2.9. Network Size) addressed this challenge by reducing the number of required channels by 79.12% (mean 16 contacts per patient) with only a modest loss in performance, supporting previous findings that reduced channel sets are sufficient for seizure prediction ^33–35^.

In patients with poor surgical outcomes, EN-based channels outperformed clinically selected resection areas for online state detection, suggesting that the estimated EN captures informative epileptogenic dynamics. While these findings support the value of reliable EN biomarkers for both surgical targeting and seizure prediction, they do not establish the EN as the optimal target, as other brain regions may also contain predictive information. Overall, the results indicate that implantation strategy influences performance and demonstrate that the proposed pipeline is compatible with current and future implantable BCI hardware.

### 4.5 General Contributors to Model Performance Variability

Classifier choice emerged as the most influential pipeline component, with RF outperforming LR and SVM, consistent with its higher modelling capacity and previous studies ^16,36^. LR also outperformed SVM, in line with some prior reports ^12^ and its competitive FPR/h performance ^29^, although opposite trends have also been described, often requiring greater tuning and higher computational cost for SVM ^37^. Beyond performance, simpler models provide advantages in efficiency and interpretability.

As no consensus exists on the optimal FC metric for preictal detection, we evaluated several commonly used approaches while excluding methods requiring manual input to preserve automation. FC metric was the second most influential component of the pipeline. Amplitude-based measures (CC and Pearson correlation) outperformed phase-based metrics (PLV, PLI) and PAC, consistent with previous studies ^38–39^, while also offering greater computational efficiency for real-time applications ^40^. GC was excluded due to inconsistent convergence, likely reflecting the non-stationary nature of SEEG signals, in agreement with previous reports questioning its suitability for seizure prediction ^41^.

Monopolar referencing achieved the best performance among the evaluated schemes. Although direct comparisons in seizure prediction are limited, monopolar configurations appear more robust to incomplete electrode sampling ^42^ and are computationally simpler, supporting their suitability for real-time applications.

Although leading eigenvector decomposition substantially reduces dimensionality and computational cost, using the full CM yielded better performance. This contrasts with previous studies reporting preserved predictive performance using alternative summarisation approaches such as graph-based methods ^43^.

Delta and gamma bands yielded the highest performance, consistent with previous reports linking these frequencies to seizure-related connectivity dynamics ^44^, brain state classification ^45^, and seizure onset zones ^46^. In particular, gamma-band connectivity has been associated with increased epileptogenic activity, especially when estimated using amplitude-based measures ^39^.

### 4.6 Clinical Implications, Trade-Offs, and Future Directions

Although recent research has increasingly focused on multi-day probabilistic seizure forecasting, patients and caregivers consistently prefer short, deterministic prediction horizons (<24 hours) ^13^ because they enable timely safety measures and administration of rescue medication. By achieving continuous prediction with a median SPH of 14 to 20 minutes, our framework demonstrates that clinically actionable short-horizon prediction remains a viable goal.

During continuous validation, the selected models achieved a false warning rate of 1.51 to 1.69 events per hour. Although this still corresponds to approximately 36 warnings per day, it may be acceptable in closed-loop neurostimulation systems, where stimulation is delivered automatically without disturbing the patient, provided sensitivity remains high. Future incorporation of multi-stage confirmation algorithms or additional physiological signals, such as heart rate variability, could further reduce false alarms.

### 4.7 Limitations

The main limitation of this study is that recordings were acquired under controlled clinical conditions, including hospital monitoring, reduced patient activity, and anti-seizure medication withdrawal, which may not fully reflect seizure dynamics during daily life. Furthermore, all patients were recruited from a single centre. Although external validation would be desirable, to our knowledge, no comparable publicly available dataset currently exists. In addition, while recordings averaged approximately 37 hours per patient, some patients experienced only two seizures, limiting validation to a single unseen event and restricting assessment of long-term generalisation.

Although the proposed framework approximates real-world deployment through continuous online training and testing, definitive proof of clinical feasibility would require prospective in vivo validation. Finally, the search and selection of validated models could be further optimised, for example by secondary selection criteria into the evolutionary selection process.

## 5. Conclusion

This study demonstrates that actionable, short-horizon seizure prediction is feasible when combined with continuous online evaluation and modern computing infrastructure. By releasing an open continuous SEEG dataset and introducing an online framework that supports iterative post-event model retraining, this work provides a foundation for the development of self-adaptive, hardware-efficient closed-loop brain-computer interfaces for seizure prediction.

## Data Availability

Abridged and anonymized data from the Hospital del Mar Seizure Prediction Dataset are available from the outset. To ensure patient privacy and data security, raw recordings are not directly distributed, and the complete set of channels per patient is accessible exclusively in a federated format stored on the Hospital del Mar - IMIM servers.
Patient-wise data may only be exported after demonstrably irreversible post-processing that prevents reconstruction or reverse engineering of the original recordings while preserving privacy and clinical context. Researchers requiring additional information (e.g., anatomical details) or wishing to work with the full dataset should consult the website (under construction) for applicable procedures.
Users are required to refer to the dataset as the "Hospital del Mar Seizure Prediction Dataset" and to cite this article in any publications resulting from its use.

## Acknowledgements

We thank the help of the nurse team of the Epilepsy Monitoring Unit of Hospital del Mar, without whom SEEG data acquisition would not have been possible. This research was funded by Agència de Gestió d’Ajuts Universitaris i de Recerca (AGAUR) Generalitat de Catalunya grant number FI-SDUR 20203.

## Author Contributions

**Justo Montoya-Gálvez**: Conceptualization (lead); data curation (lead); formal analysis (lead); funding acquisition (supporting); investigation (lead); methodology (lead); software (lead); validation (lead); visualization (lead); writing – original draft preparation (lead); writing – review and editing (lead). **Karla Ivankovic**: Investigation (supporting); software (supporting); supervision (supporting); writing – review and editing (supporting). **Marizeh Nazari**: Data curation (supporting); software (supporting). **Alessandro Principe**: Conceptualization (lead); data curation (lead); funding acquisition (lead); methodology (supporting); project administration (lead); resources (lead); supervision (lead); visualization (supporting); writing – original draft preparation (supporting); writing – review and editing (lead). **Rodrigo Rocamora**: Resources (lead); funding acquisition (lead); supervision (lead); writing – original draft preparation (supporting); writing – review and editing (lead).

## Conflict of Interest/Ethical Publication Statement

None of the authors has any conflict of interest to disclose. We confirm that we have read the Journal’s position on issues involved in ethical publication and affirm that this report is consistent with those guidelines.

