## Supplemental Files for "Countering Neural Activity Drift: Sustained Long-term Seizure Prediction Using an Evolutionary Machine-Learning Framework on Continuous Intracranial EEG"

#### Table S1: Inclusion Criteria

A total of 50 patients were initially assessed. Exclusion occurred due to: no surgery (n = 22), palliative procedures (n = 2), insufficient follow-up (n = 3), fewer than two recorded seizures (n = 6), and inaccessible recordings (n = 1). This resulted in 16 included patients, of whom 12 achieved good surgical outcome (Engel I) and 4 had poor outcome (Engel II–IV).

| Criterion | Number of patients (n) |
|---|---|
| Admitted for presurgical evaluation | 50 |
| Did NOT proceed to surgery | 22 |
| Underwent palliative surgical interventions (e.g. vagus nerve stimulation, callostomy, neuromodulation) | 2 |
| Did NOT meet the required minimum follow-up duration | 3 |
| Did NOT exhibit at least the required minimum spontaneous seizures during presurgical SEEG monitoring | 6 |
| Not possible to open recordings | 1 |
| **Total included in the study** | **16** |
| Good outcome (Engel I) | 12 |
| Bad outcome (Engel II-IV) | 4 |

#### Table S2: Dataset Benchmarking

| Dataset | Cohort size | Number of channels | Total recording duration | Continuous data |
|---|---|---|---|---|
| Presented | 16 | 104.65 [27-150] | 664.9h | Yes |
| Melbourne NeuroVista seizure trial<br><br>Brinkmann 2016 | 12 | 16 | Around 88h | No |
| University of Bonn | 10 | 1 | 3.19h | No |
| Kaggle UPenn and Mayo Clinic's Seizure Detection | 12 | 16-76 | 7,2h | No |

| | | | | |
|---|---|---|---|---|
| Challenge<br><br>Temko 2015 | | | | |
| Kaggle American Epilepsy Society Seizure Prediction Challenge | 7 | 16 | 677,84h | No |

#### Table S3: Welch's ANOVA

Welch's ANOVA was applied due to violations of normality and homoscedasticity assumptions across all factors. The full results are shown below:

| Source | F | p-value | Partial \eta_p^2 |
|---|---|---|---|
| Classifier | 20785.548 | <0.001 | 0.270 |
| Connectivity Method | 2259.0394 | <0.001 | 0.045 |
| Target area | 654.466 | <0.001 | 0.004 |
| Reference | 130.840 | <0.001 | 0.002 |
| Feature | 124.898 | <0.001 | 0.001 |
| Band | 36.056 | <0.001 | 0.002 |

All factors showed statistically significant main effects (all p < 0.001). Effect sizes were computed as partial eta squared (\eta_p^2), indicating the proportion of variance in AUROC explained by each factor within the model.

#### Table S4: Games-Howell Summary

Full Games–Howell post hoc results are summarised in the table below. All pairwise comparisons were statistically significant (p < 0.001).

| **Pipeline Step** | **Best Method** | **Mean AUROC** | **Difference with Runner-up** |
|---|---|---|---|
| Classifier | Random Forest | 0.944 | +0.089 |
| Connectivity | Cross Correlation | 0.887 | +0.088 |
| Network | Full Implantation | 0.818 | +0.039 |
| Reference | Monopolar | 0.816 | +0.025 |
| Feature representation | Full Connectivity Matrix | 0.807 | +0.017 |
| Band | Delta | 0.817 | +0.022 |

Pairwise comparisons were performed using the Games–Howell test following Welch’s ANOVA to account for unequal variances and sample sizes.

### Figure S1: Comparison between sample-based and event-based evaluation of online classifier performance

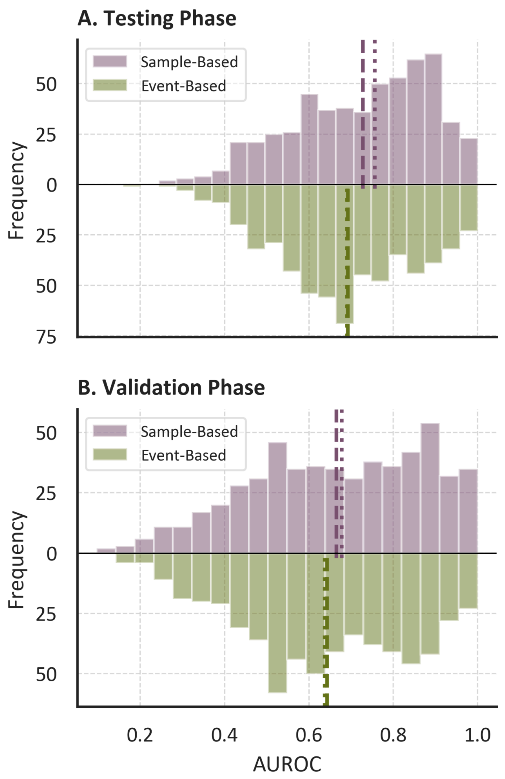


**(A)** Mirrored histogram showing the distribution of area under the receiver operating characteristic curve (AUROC) values during the testing phase for classifiers achieving full-prediction of all seizures. Light blue histograms represent sample-based evaluation, in which performance was assessed at the epoch level, whereas dark blue histograms represent event-based evaluation, in which seizures were considered correctly predicted regardless of potential mismatches between interictal and preictal epoch labels. Dashed lines indicate the mean of each distribution, and dotted lines indicate the median. **(B)** Mirrored histogram showing the distribution of AUROC values during the validation phase for classifiers achieving full-prediction. Light blue histograms correspond to sample-based evaluation and dark blue histograms correspond to event-based evaluation. Dashed lines indicate the mean of each distribution, and dotted lines indicate the median.

### Appendix S1: Electrodes

Electrodes consisted of 0.8 mm diameter depth contacts with 5–15 contacts per shaft (2 mm length, 1.5 mm inter-contact spacing). Missing signal segments were replaced with zeros to maintain temporal alignment across recordings. Selection of usable data prioritized segments preceding clinical stimulation procedures.

### Appendix S2: Epoching

The 60-second epochs were extracted using a sliding window with a 30-second step. However, to reduce edge artefacts from filtering, 80-second segments were initially extracted (60-second epochs with 10-second padding margins on each side) to minimize edge artifact, although only the central 60 seconds were used for subsequent functional connectivity analysis.

### Appendix S3: Referencing

Three referencing montages were evaluated independently: monopolar, bipolar, and Laplacian.

In monopolar montage each channel reflects the potential difference between an intracerebral contact and a common extracerebral reference.

In the bipolar montage, each signal represents the potential difference between adjacent contacts along the same electrode shaft. For an electrode $e$ with $n$ contacts, the bipolar signals $\left\{s^e_{i,(i+1)} \mid i =1,2, \ldots,n-1 \right\}$ are derived from the monopolar signals $\left\{s^e_i \mid i =1,2, \ldots,n \right\}$ as:

\begin{equation}
\label{eq:BipolarReference}
s^{e}{i,(i+1)} = s^{e}{i} - s^{e}_{i+1}, \qquad i = 1, 2, \ldots, n-1.
\end{equation}

The Laplacian montage approximates the second spatial derivative of the potential, enhancing local activity while attenuating common-mode components (Appendix S1). For the same electrode $e$ with $n$ contacts, the Laplacian-referenced signals $\left\{s'^e_i \mid i =1,2, \ldots,n \right\}$ were computed as:

\begin{equation}
\label{eq:LaplacianReference}
\left\{
\begin{array}{ll}
s'^e_{i} = s^e_{i} - s^e_{i+1} & \text{for } i = 1 \\
s'^e_{i} = s^e_{i} - \dfrac{s^e_{i-1} + s^e_{i+1}}{2} & \text{for } i = 2, \ldots, n-1 \\
s'^e_{i} = s^e_{i} - s^e_{i-1} & \text{for } i = n
\end{array}
\right.
\end{equation}

### Appendix S4: Artifacts Removal

To attenuate power-line interference, a notch (band-stop) filter was applied at the fundamental frequency $f_C=50$ Hz and its harmonics. A stopband width of 5 Hz was used. The filter quality factor $Q$ was computed as

\begin{equation}
\label{eq:QfactorNotch}
Q= \frac{f_C}{f_H - f_L}
\end{equation},

where $f_H$ and $f_L$ denote the upper and lower bounds of the stopband, respectively.

A digital Butterworth high-pass filter with a cut-off frequency of 0.1 Hz was applied to remove slow drifts and baseline fluctuations that could obscure physiologically meaningful neural oscillations. Following filtering, only the central 60 seconds of each 80-second epoch were retained, as previously described.

### Appendix S5: Classifiers

Each connectivity matrix was serialised and used as input to a supervised machine learning pipeline.

Three common supervised classifiers in the field of epilepsy with increasing model complexity were evaluated: (1) a regularised Logistic Regression (LR), (2) a Support Vector Machine (SVM), and (3) a Random Forest (RF) classifier.

The RF classifier was trained with class balancing, where class weights were set inversely proportional to class frequencies according to Eq.~\ref{eq:class_imbalacing}, thereby mitigating bias toward the dominant interictal class:

\begin{equation}
w_c=\frac{n_{\text{samples}}}{n_{\text{classes}}\times\text{bincount}(y)}.
\label{eq:class_imbalacing}
\end{equation}

The SVM employed a radial basis function (RBF) kernel, with the kernel coefficient $\gamma$ set to $1/n_{\text{features}}$, where $n_{\text{features}}$ corresponds to the number of pairwise connectivity features. The classifier was configured to output calibrated probabilities of belonging to the preictal class.

Three reference classifiers were included as baselines: (4) a Majority Class Classifier (MCC) that always classifies the state as interictal, (5) a Minority Class Classifier (MinCC) that always classifies the state as preictal, and (6) a Random-Stratified (STR) classifier, which assigns labels randomly while preserving the original interictal-to-preictal class proportions.

### Appendix S6: Data Augmentation

The recording durations ranged from 15 hours to 3 days per patient, which was insufficient to adequately evaluate the classification algorithms under conditions representative of real-world deployment. To address this limitation, the dataset was artificially expanded through repeated resampling. The continuous signals were segmented into 60-second epochs, consistent with the configuration described previously, while the overlap between consecutive epochs was randomly selected for each epoch from the integer interval [15, 45] seconds. This resampling procedure was repeated 1000 times.

Epochs were labelled as interictal, preictal, ictal, and subclinical as described in previous sections. Due to the consecutive sampling and the random temporal shifts introduced by this procedure, some epochs encompassed the precise transition to the ictal state. These epochs were labelled as transition.

### Appendix S7: Online Testing

The evaluation of each classifier was divided into two phases: (1) A pseudo-prospective leave-one-seizure-out testing phase, and (2) a pseudo-prospective validation phase under realistic class imbalance.

In the first phase, a synthetic continuous signal was generated by concatenating the interictal, preictal, and ictal segments corresponding to the seizure used for training together with the remaining seizures, excluding one randomly selected seizure (distinct from the one used for training), which was reserved for the second phase of testing. It is important to note that epoch segmentation during training incorporated a random temporal offset, whereas in the testing phases the classifier operated using a sliding window of 60 seconds duration with a step size of 30 seconds. Consequently, even when the classifier encountered the same seizure used during training, the resulting connectivity matrices were not necessarily identical, as the precise temporal boundaries of the epochs could differ. The order in which seizures were presented was randomised, while preserving the chronological relationship between the interictal and preictal intervals preceding each seizure. More specifically, the signal was partitioned into blocks spanning from the end of one seizure to the end of the subsequent seizure. These blocks were then randomly reordered, concatenated, and treated as a continuous recording.

This procedure led to two particular cases. First, the interictal segment preceding the first recorded seizure is not preceded by any seizure; in this case, the beginning of the recording was considered the start of the block. Second, the interictal segment following the final seizure is not followed by another seizure; this block was reserved for the validation phase of the testing, as described below.

Once the artificial continuous signals were constructed, the classifiers were evaluated in an online manner, processing the signal epoch by epoch. Each CM was classified as preictal when the probability estimated by the RF exceeded 50%, and as interictal otherwise. When an epoch was classified as preictal, the subsequent 5 minutes of signal were automatically labelled as preictal, and their associated probabilities were not updated unless a higher probability value was obtained. If the estimated probability of the preictal state remained stable or increased, the duration of the detected preictal period could extend beyond this 5-minute interval.

Performance was first evaluated at the sample level, where each epoch was treated as an individual sample. The computed metrics included recall, the warning time defined as the total number of epochs classified as preictal divided by the recording duration (expressed as preictal samples per hour), the false warning time defined as the number of incorrectly classified preictal epochs divided by the recording duration (expressed as false positive samples per hour), and AUROC.

However, this approach assumes a fixed preictal duration of 5 minutes, which was originally selected based on patient preferences and may not reflect the true temporal extent of the preictal state. To account for potentially longer preictal dynamics, performance metrics were also computed at the event level. Consecutive preictal detections without interruption were considered as a single preictal event. If the end of a detected preictal event coincided with seizure onset, the event was considered a true positive. Conversely, if the detected preictal event terminated before seizure onset, it was considered a false positive. Recall, warning time, and false warning time were recomputed at the event level. Additionally, for correctly detected preictal events, the SPH was calculated as the time interval between the onset of the detected preictal state and seizure onset.

Following an evolutionary algorithm-inspired framework, only classifiers achieving an event-level recall of 1.0 were retained for the validation phase. In other words, only models that successfully predicted all seizures in the initial phase were advanced to the second stage.

In this second phase, the block corresponding to the previously excluded seizure was used, followed by concatenation with the block containing only interictal activity after the final recorded seizure. This procedure aimed to increase the amount of interictal data and more closely approximate the class imbalance observed in clinical recordings. The same performance metrics computed in the testing phase were also evaluated during this validation phase. As before, only models achieving complete recall were considered successful. Consequently, models that passed both the testing and validation stages were able to predict all seizures, although the quality of the predictions, in terms of warning time, number of events, and SPH, could vary.

### Appendix S8: Subjects and Recordings (Results)

The good outcome group was defined as patients achieving complete seizure freedom (Engel Class I) at last follow-up (3 years after resective surgery). The bad outcome group included patients with Engel Classes II–IV.

Mean age, standard deviation, and range for each subgroup were computed as reported. Channel-level statistics included total number of SEEG contacts per patient, mean ± SD, and range across the cohort.

Seizure counts were computed per patient and aggregated by group. Mean, median, standard deviation, and range of seizure counts were reported across the cohort and within each outcome group.

The dataset size comparison with existing intracranial EEG datasets was based on reported total recording durations and cohort sizes from previously published datasets, including the Kaggle American Epilepsy Society Seizure Prediction Challenge dataset (677.84 h).

The assumption that EN identification is reliable only in patients with good surgical outcomes is based on the premise that seizure freedom indicates adequate sampling and resection of the epileptogenic network.

### Appendix S9: Reference, Connectivity Representation, and Frequency Effect (Results)

Effect sizes for reference type, connectivity matrix reduction, and band filtering were small ($\eta_p^2 \approx 0.01$).

**Referencing schemes:**

- Monopolar: AUROC = 0.816
- Bipolar: AUROC = 0.790
- Laplacian: AUROC = 0.791
- Bipolar vs Laplacian: $p = 0.977$ (not significant)
- Monopolar vs bipolar and Laplacian: $p < 0.001$

**Connectivity representation:**

- Full connectivity matrix: AUROC = 0.807
- Eigenvector-based reduction: AUROC = 0.791
- Difference: $p < 0.001$

**Frequency band results:**

- Delta: AUROC = 0.817
- Theta: AUROC = 0.808 ($p < 0.01$ vs delta)
- Alpha: AUROC = 0.800 (no significant difference vs most bands, $p > 0.05$, except delta $p < 0.001$)
- Low gamma: AUROC = 0.795 ($p < 0.001$)
- High gamma: AUROC = 0.796 ($p < 0.001$)
- Broadband: AUROC = 0.778 ($p < 0.001$)

These comparisons indicate limited but measurable modulation of performance by preprocessing choices, with effects substantially smaller than those observed for classifier type and connectivity metric selection.

### Appendix S10: Functional Connectivity Measurements (Results)

All feature combinations were evaluated except those involving GC which frequently failed to converge. The remaining configurations yielded 1,263 feature pipelines and 12,630 trained classifiers, enabling a large-scale comparison of preprocessing choices in SEEG-based seizure prediction.
